# Application of AMR in evaluating microvascular dysfunction after ST-segment elevation myocardial infarction

**DOI:** 10.1101/2023.05.11.23289795

**Authors:** Hao Wang, Qi Wu, Lang Yang, Long Chen, Wen-Zhong Liu, Jing-Song Xu

**Author notes:** Corresponding author: Department of Cardiology, The Second Affiliated Hospital of Nanchang University, 1 Minde Road, Nanchang 330006, China.

## Abstract

**BACKGROUND:** Quantitative flow ratio (QFR) is a new method to estimate fractional flow reserve based on three-dimensional quantitative coronary angiography, from which angiography-derived microcirculatory resistance (AMR) without guidewires and adenosine is derived as an indicator of microvascular dysfunction. This study aimed to assess coronary microvascular dysfunction (CMD) in patients with ST-segment elevation myocardial infarction (STEMI) by AMR.

**METHODS:** A retrospective collection of 506 STEMI patients who successfully underwent percutaneous coronary intervention (PCI) from June 1, 2020, to September 28, 2021, was divided into the CMD group and the non-CMD group based on the value of AMR, while we used propensity score matching (PSM) to adjust for baseline characteristics. The primary endpoint was the 1-year rate of major adverse cardiac events (MACE), a composite of death from any cause, myocardial infarction, readmission for heart failure, or ischemia-driven revascularization.

**RESULTS:** The 1-year rate of MACE in CMD group was higher than that in the non-CMD group (post-match HR 1.954, 95% CI:1.025 to 3.726; 14.1% vs. 7.3%, P=0.042); Subgroup analysis showed that the readmission rate of heart failure (HF) was higher in the CMD group than in the non-CMD group (post-match HR 5.082, 95% CI:1.471 to 17.554; 7.9% vs. 1.6%. P=0.010). The results of survival analysis suggested that AMR ≥250mmHg*s/m was an independent predictor of the primary endpoint in STEMI patients (post-match adjusted HR 2.265, 95% CI: 1.136 to 4.515, P = 0.020). CONCLUSION: As an indicator of microvascular dysfunction, AMR can be a viable alternative to invasive wire-based IMR in STEMI patients.

## 1. Introduction

Coronary microvascular dysfunction (CMD) plays an important role in myocardial ischemia in many cardiovascular diseases, and coronary artery disease (CAD) is a common cardiovascular condition in which CMD can contribute to its formation and progression [1,2]. Notably, post-reperfusion CMD is not uncommon in ST-segment elevation myocardial infarction (STEMI) patients, and the occurrence of microvascular obstruction (MVO) is also associated with poor clinical prognosis [3].

The index of microcirculatory resistance (IMR) is a quantitative and repeatable method for evaluating CMD based on intracoronary guidewire [4], but the clinical application of IMR is still limited by dilating drugs, pressure guidewires, long measurement times, and high costs. Functional assessment is becoming increasingly important in patients with coronary artery disease, and quantitative flow ratio (QFR) is an emerging angiography-based assessment method that facilitates the calculation of fractional flow reserve (FFR) values by three-dimensional (3D) coronary reconstruction and fluid dynamics [5]. The accuracy of QFR has been confirmed by many large RCT studies, including the FAVOR Pilot study, the FAVOR II study, and the FAVOR III study, which further validated the diagnostic value of QFR and the accuracy of assessing coronary ischemia [5–10]. In addition, QFR eliminates the need for pressure guidewires and is a suitable option for assessing coronary ischemia [6]. And a guidewire-free and adenosine-free angiography-derived microcirculatory resistance (AMR) derived from QFR with flow velocity calculation has good diagnostic accuracy in the assessment of CMD, providing an effective clinical alternative to invasive pressure guidewire-based IMR [11].

Although QFR is an important tool for functional assessment in patients with CAD, the specific application and assessment value of AMR in STEMI patients still lacks support from relevant studies. Therefore, our study aimed to explore the value of AMR in assessing the incidence of major adverse cardiac events (MACE) by evaluating postoperative AMR and follow-up characteristics in STEMI patients.

## 2. Materials and Methods

### 2.1. Study Design

This study was a single-center observational study, which included STEMI patients who successfully underwent percutaneous coronary intervention (PCI) at The Second Affiliated Hospital of Nanchang University from 2020.06.01 to 2021.09.28. All enrolled patients were evaluated noninvasively by AMR for the microcirculatory status of the culprit vessel, and postoperative AMR, QFR, and 1-year follow-up data were collected.

Patients were divided into the CMD group (AMR ≥ 250 mmHg*s/m, n=215) and the non-CMD group (AMR < 250 mmHg*s/m, n=291) based on a postoperative AMR value of 250 mmHg*s/m (due to the lack of established AMR cut-off values for CMD, relevant data from recent studies were used [11]), with 191 cases in both the CMD group and non-CMD group after matching.

### 2.2. Patient population

Adult patients with STEMI who underwent PCI within 12 hours of symptom onset had ≥50% stenosis of the lesion diameter on initial angiography. In the case of STEMI undergoing primary PCI, a coronary physiological assessment of the culprit vessel was performed after thrombus aspiration and/or balloon dilatation flow restoration and/or completion of primary PCI.STEMI was defined as the occurrence of persistent chest pain for at least 30 minutes with ST-segment elevation >2 mm in at least two contiguous leads or a new left bundle branch block [12]. Identification of the culprit vessel was based on (1) angiographic presentation matching the presence of plaque instability or thrombus, and (2) electrocardiographic and echocardiographic findings. Two angiographic images with at least 25° projection angle separation were measured and the data were transferred to the AngioPlus system (Pulse Medical Imaging Technology, Shanghai, China) via the local site network for QFR, AMR calculation. Patients were further excluded if QFR and AMR could not be calculated: (1) poor quality of angiographic images; (2) presence of severe vascular curvature, and overlap [7] [13].

The retrospective study was approved by the Institutional Review Boards at The Second Affiliated Hospital of Nanchang University, Nanchang, China, which conforms to the declaration of Helsinki. The data are anonymous, and the requirement for informed consent was therefore waived.

### 2.3 Data collection

The following parameters were retrospectively collected using medical records: age, gender, cigarette smoking, history of AMI or PCI, and clinical comorbidities including hypertension, diabetes, and hyperlipidemia. Serum biochemical markers such as glucose, low-density lipoprotein (LDL-c), B-type natriuretic peptide (BNP), C-reactive protein (CRP), creatinine, and troponin I were measured in the hospital clinical laboratory using routine automated techniques.

### 2.4 QFR and AMR Analysis

The AMR and QFR calculations in this study were performed by the AngioPlus system (Pulse Medical Imaging Technology, Shanghai, China) according to standard operating procedures.

Anatomical information of the target vessel, including lumen diameter and lesion length, was provided by revascularization. An end-diastolic frame was selected for each projection and the images preferably had frames from the same cardiac cycle. The reference vessel was constructed by the system on a healthy segment, ideally located proximal and distal to the target lesion. AMR was calculated by the Angioplus System, as shown in Figure 2.

### 2.5 Clinical Follow-up

Relevant clinical data and 1-year major adverse cardiac events (MACE) were recorded for all enrolled individuals during their hospitalization. MACE were defined as a composite of death from any cause, any myocardial infarction (MI), readmission for heart failure, or any ischemia-driven revascularization. All patients were treated according to the clinical guidelines recommended at the time of discharge. The occurrence of MACE within one year was documented by telephone follow-up and review of medical records.

Cardiac-related death was defined as death due to myocardial infarction, severe arrhythmia, refractory heart failure, or cardiogenic shock. Readmission for heart failure was defined as hospitalization due to new or worsening signs and symptoms of heart failure with concurrent noninvasive imaging findings or increased BNP concentrations and discharge with a diagnosis of congestive heart failure. Spontaneous myocardial infarction was defined as elevated creatine kinase or troponin levels above the upper limit of normal with symptoms of ischemia or ECG findings suggestive of ischemia [14]. Ischemia-driven target revascularization was defined as revascularization with at least one of the following:(1) recurrence of angina pectoris; (2) positive noninvasive test; and (3) positive invasive physiological test.

### 2.6 Statistical analysis

Continuous variables were recorded as mean ± standard deviation (SD) or median (25th percentile, 75th percentile), and categorical variables were recorded as counts (percentages). Normality was tested appropriately with the Kolmogorov-Smirnov test or the Shapiro-Wilk test. Comparisons between categorical variables were assessed using Pearson Chi-squared test or Fisher’s exact test (as appropriate). To reduce potential bias between groups, we conducted propensity-score matching (PSM) to balance baseline characteristics. For the PSM, the 1:1 nearest neighbor matching was used without replacement and a caliper of 0.02. The time-to-first event rates were estimated for each group using Kaplan-Meier methods and were compared by the log-rank test. Between-group differences were estimated by hazard ratios (HRs) with 95% CIs using a Cox proportional hazards model. Clinically relevant covariates or univariate variables associated with outcome (P < 0.10) were entered into the multivariate Cox model. Included covariates included age, male, body mass index, hypertension, diabetes, hyperlipidemia, Previous stroke, HBA1C, BNP, albumin, peak troponin I, random blood glucose, LVEF, and AMR ≥250 mmHg*s/m. p values < 0.05 were considered significant, and all comparisons were two-sided. All analyses were performed using SPSS 26.0 (IBM Inc., New York, NY, USA).

## 3. RESULTS

### 3.1 Baseline characteristics

As shown in Figure 1, this study included a total of 514 patients with AMI, 3 patients refused PCI procedures, and 5 patients had poor quality QFR images, resulting in the inclusion of 506 STEMI patients divided into the CMD group (AMR ≥250 mmHg*s/m, n=215) and the Non-CMD group (AMR <250 mmHg*s/m, n=291). The clinical, laboratory, and angiographic characteristics of each group are shown in Table 1. The mean age was 63 years, 416 males (82.2%), 90 females (17.8%), 137 (27.1%) had diabetes mellitus, 265 (52.4%) had hypertension, and 138 (27.3%) had hyperlipidemia. There was no significant difference between the CMD group and the non-CMD group in the incidence of hypertension, diabetes mellitus, previous smoking, family history of coronary heart disease, previous stroke, previous myocardial infarction, used antiplatelet drug, used statin, and LVEF (P > 0.05). Compared with non-CMD patients, CMD patients were significantly older [67.00 (56.00-74.00) vs. 60.00 (51.75-72.00) years, P=0.000], less proportionally male [167 (77.7%) vs. 249 (85.6%), P=0.022], had a lower body mass index [22.64 (20.28-23.66) vs. 22.64 (20.28-25.35) kg/m2, P=0.017], a lower proportion of hyperlipidemia [47 (21.9%) vs. 91 (31.3%), P=0.019], and less use of ACEI/ARB medications [120 (55.8%) vs. 192 (66%), P= 0.020]. Among biochemical indicators, compared with non-CMD patients, CMD patients had lower serum albumin [37.99±3.97 vs. 39.01±3.90 g/L, P=0.004], higher neutrophils [8.09 (5.79-10.46) vs. 7.16 (5.45-9.68)*10^9/L, P=0.034], and lower lymphocytes [1.41 (0.95-1.94) vs. 1.59 (1.04-2.37)*10^9/L, P=0.024], higher BNP [189.19 (78.09-485.36) vs. 142.55 (50-401.82) pg/ml, P=0.011], lower LDL-c [2.67 (2.14-3.31) vs. 2.85 (2.36-3.44) mmol/L, P=0.022]; There was no significant difference in other indexes between the two groups (P > 0.05).

**Figure 1:**
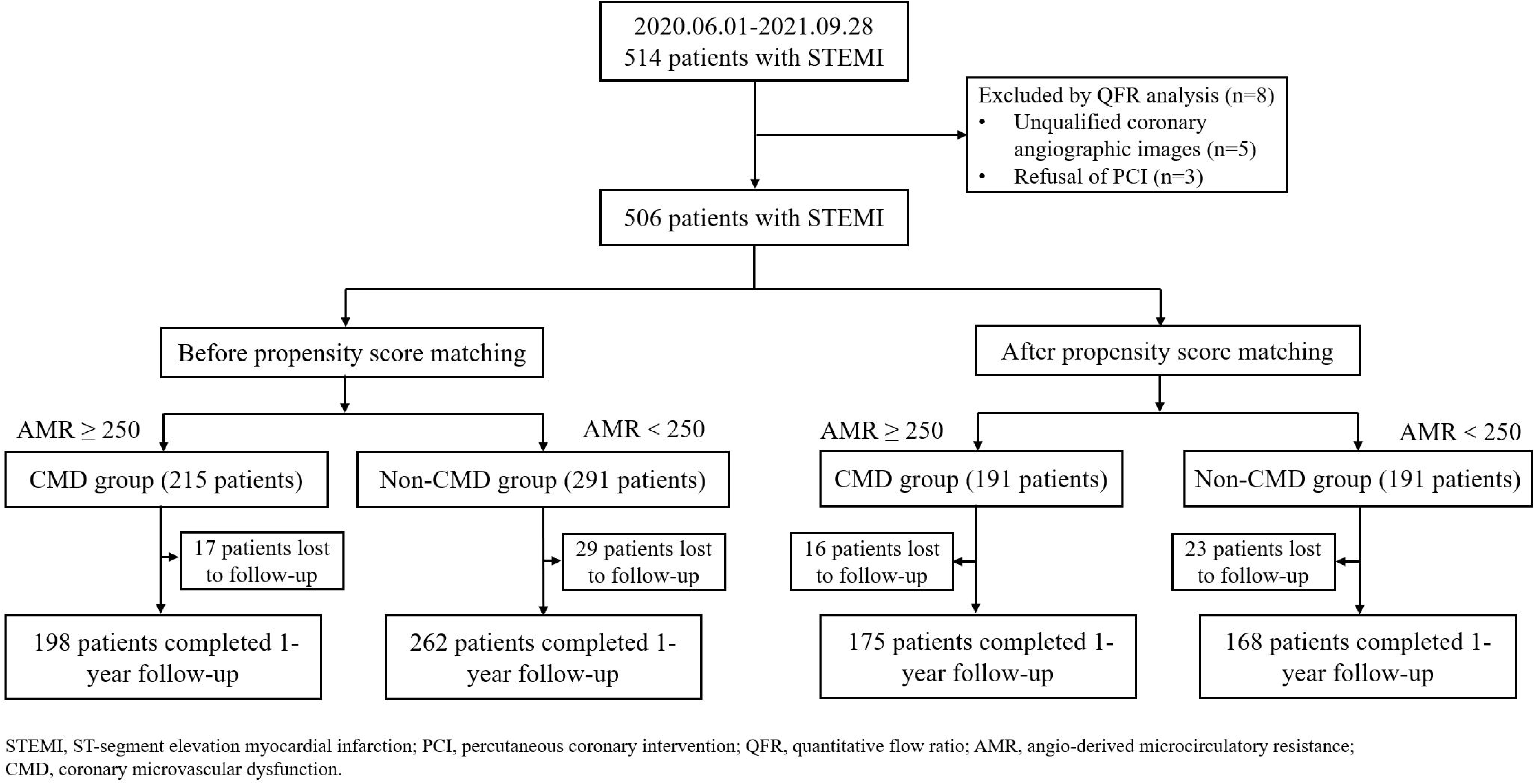
Study flowchart diagram.

**Figure 2:**
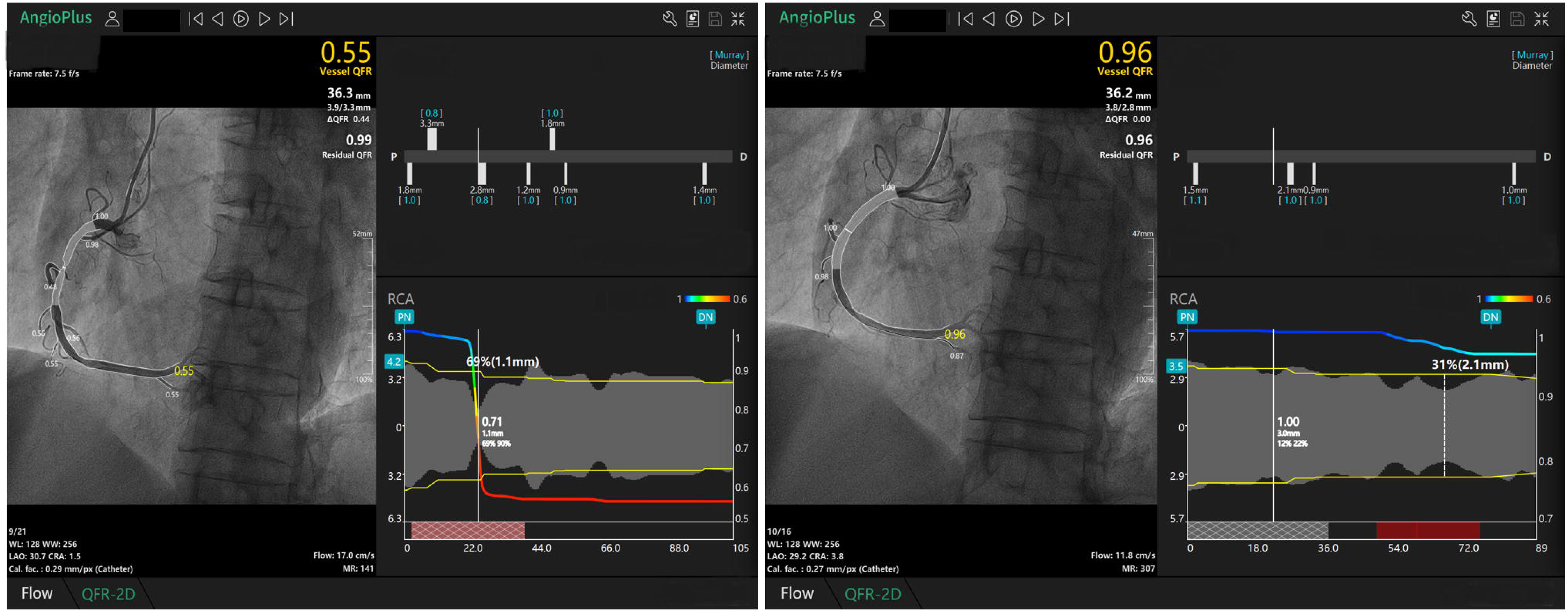
An example of Quantitative flow ratio (QFR) and Coronary microvascular dysfunction (CMD) analysis. (a) diagnostic angiographic projections show Right coronary artery (RCA) with residual stenosis; (b) QFR and AMR after PCI were calculated. QFR is 0.96, AMR is 307mmHg*s/m, flow speed is 1l.8cm/s.

**Table 1:**
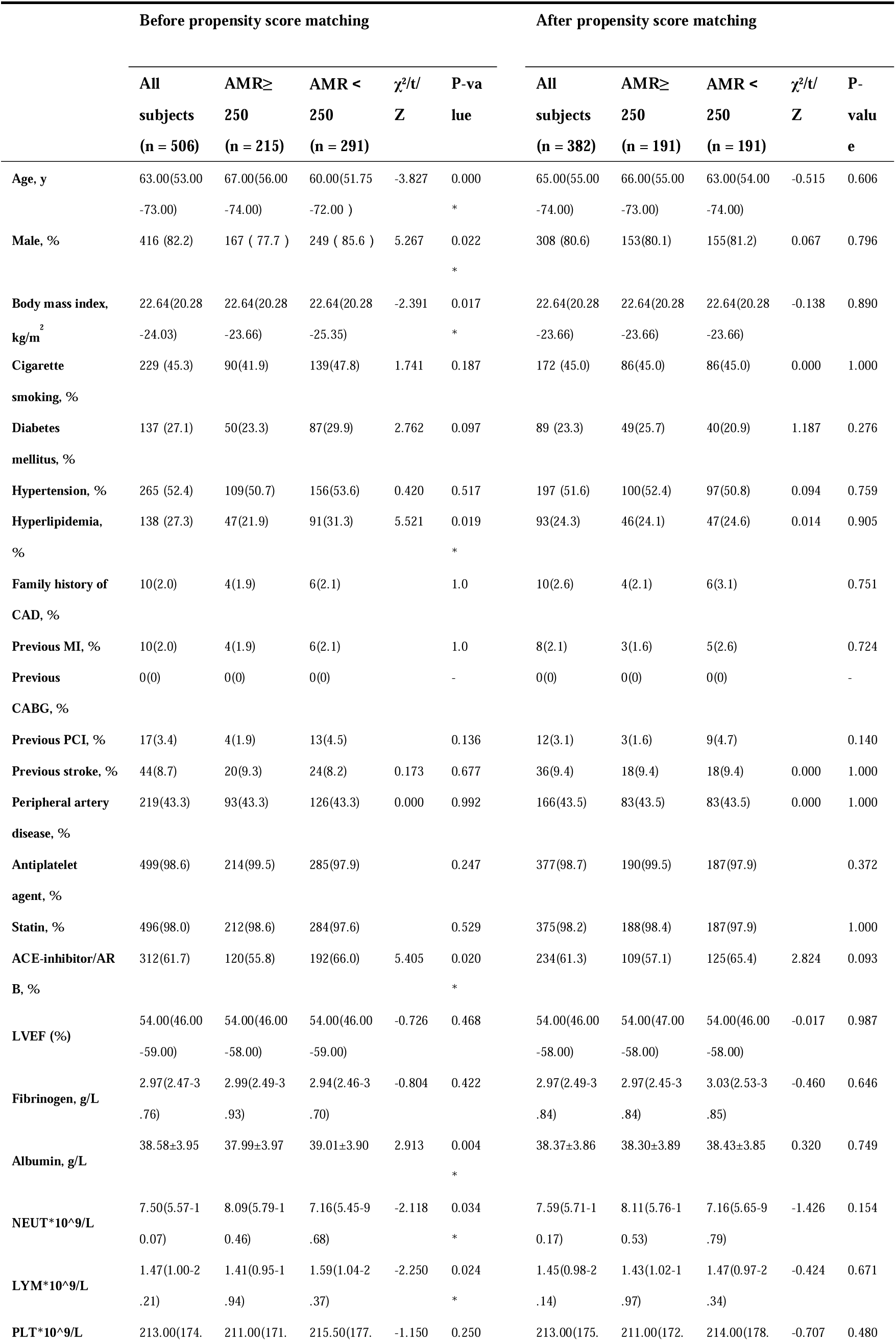
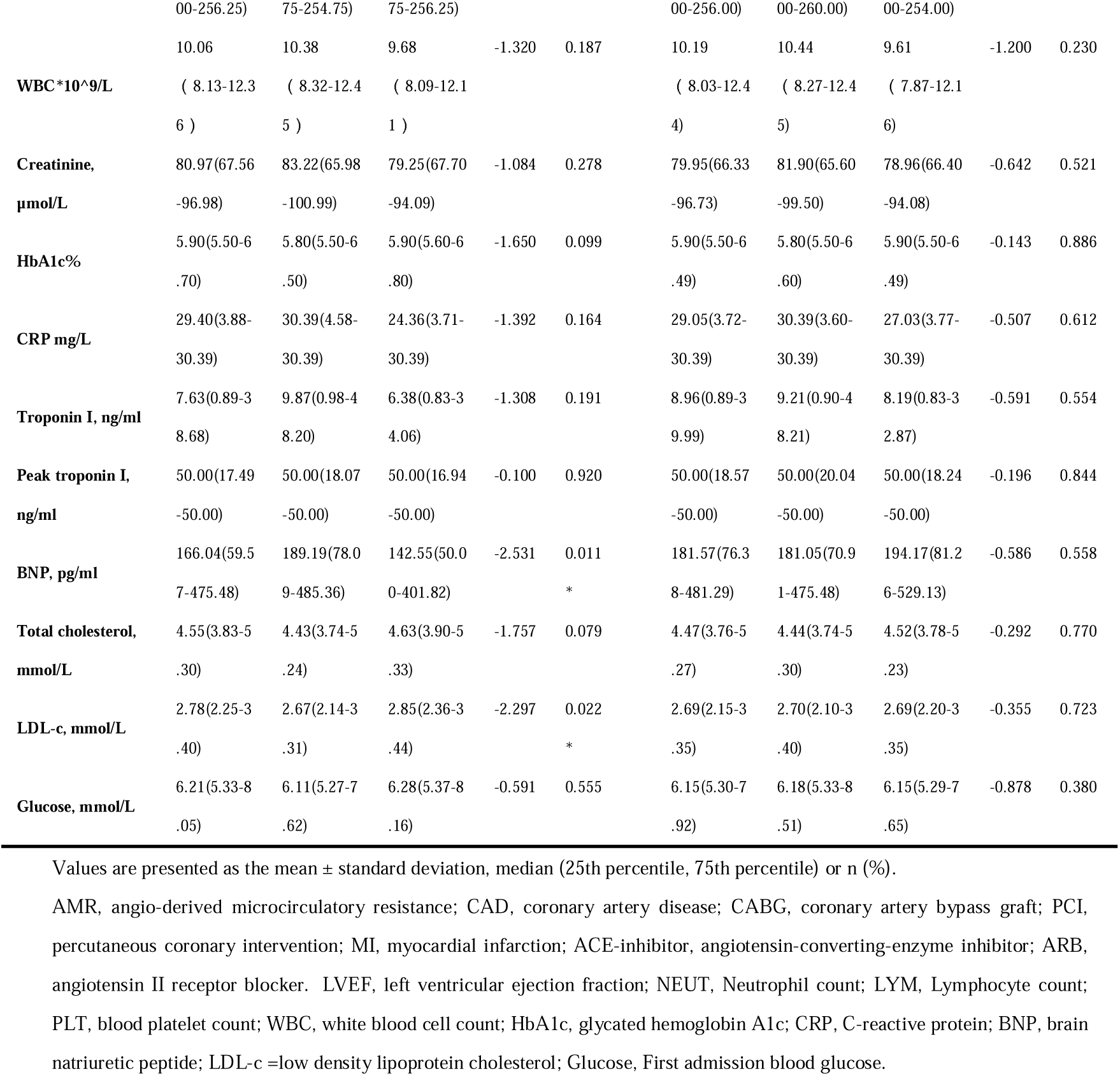
Baseline characteristics

Using the propensity score matching method (PSM) with a caliper value of 0.02, a total of 191 pairs were successfully matched between the two groups. Covariates that were not balanced between the two groups were balanced after matching (P > 0.05, Table 1).

### 3.2 Coronary angiography characteristics

Compared with non-CMD patients, the proportion of postoperative TIMI blood flow grade 3 was lower in CMD patients [206 (95.8%) vs. 291 (100%), P=0.000; 184 (96.3%) vs. 375 (98.2%), P=0.015]. In addition, pre-match and post-match radial artery routing, used bivalirudin, thrombus aspiration, multi-vessel disease, balloon pre-dilation, balloon post-dilation, Culprit vessel, number of stents, use of drug-eluting stents, drug-eluting balloons, and non-drug balloons were similarly distributed among groups (P > 0.05, Table 2).

**Table 2:**
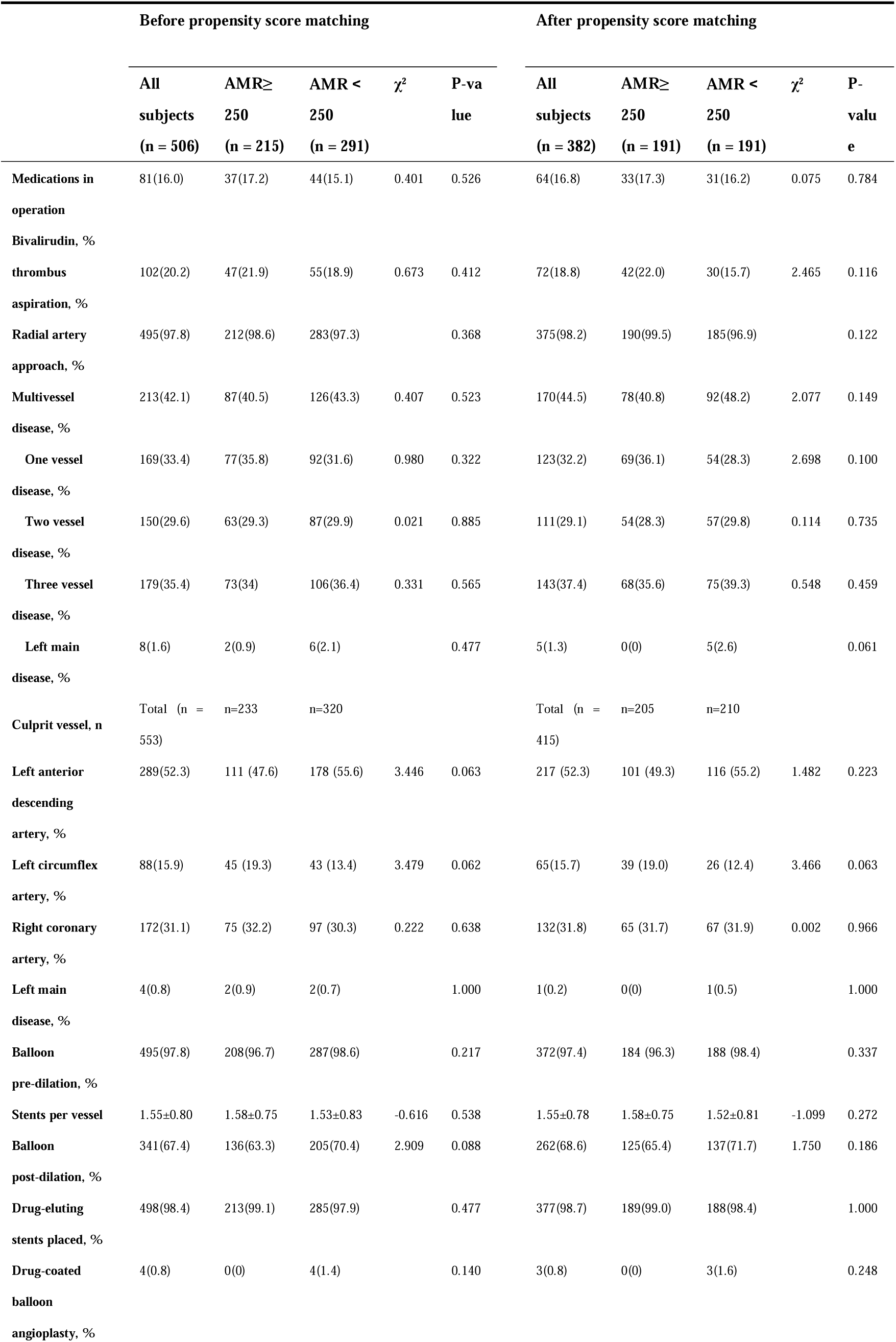
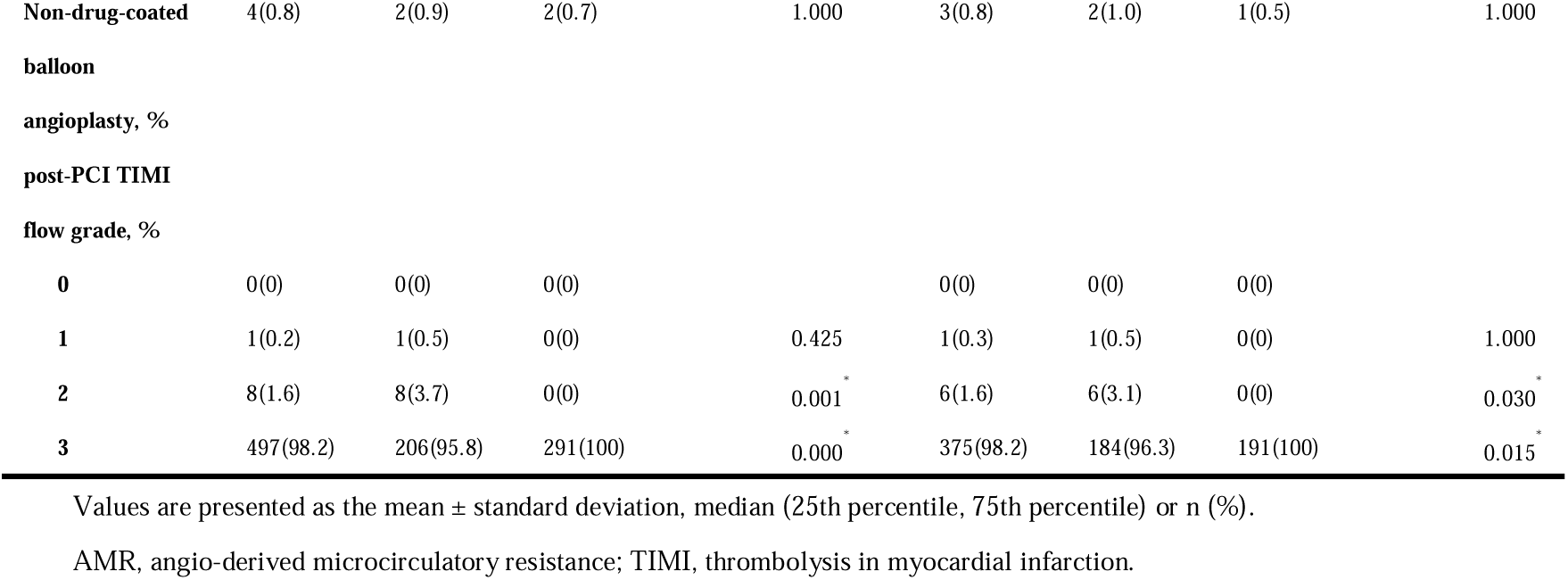
Coronary angiography characteristics

### 3.3 AMR and QFR Analysis Results

All AMR and QFR analysis data are summarized in Table 3. Compared with non-CMD patients, Post-PCI QFR was significantly higher in CMD patients [pre-match 0.97 (0.95-0.99) vs. 0.93 (0.89-0.97), P=0.000; post-match 0.97 (0.95-0.99) vs. 0.92 (0.87-0.96), P=0.000], Post-PCI AMR was significantly higher in CMD patients [pre-match 287.50 (269.00-315.50) vs. 212.00 (190.00-235.00) mmHg*s/m, P=0.000; post-match 288.00 (269.00-315.00) vs. 198.00 (183.00-222.00) mmHg*s/m, P=0.000], and Post-PCI flow velocity was significantly lower in CMD patients [pre-match 13.05 (11.10-14.60) vs. 19.8 (17.80-23.40) cm/s, P=0.000; post-match 13.10 (11.10-14.60) vs. 21.4 (18.60-25.00) cm/s, P=0.000]. In contrast, there were no statistical differences between groups in reference vessel diameter, minimum lumen diameter, stenosis diameter, lesion length, total stent length per vessel, in-stent reference vessel diameter, in-stent minimum lumen diameter, and in-stent stenosis diameter before and after matching (P > 0.05, Table 3).

**Table 3:**
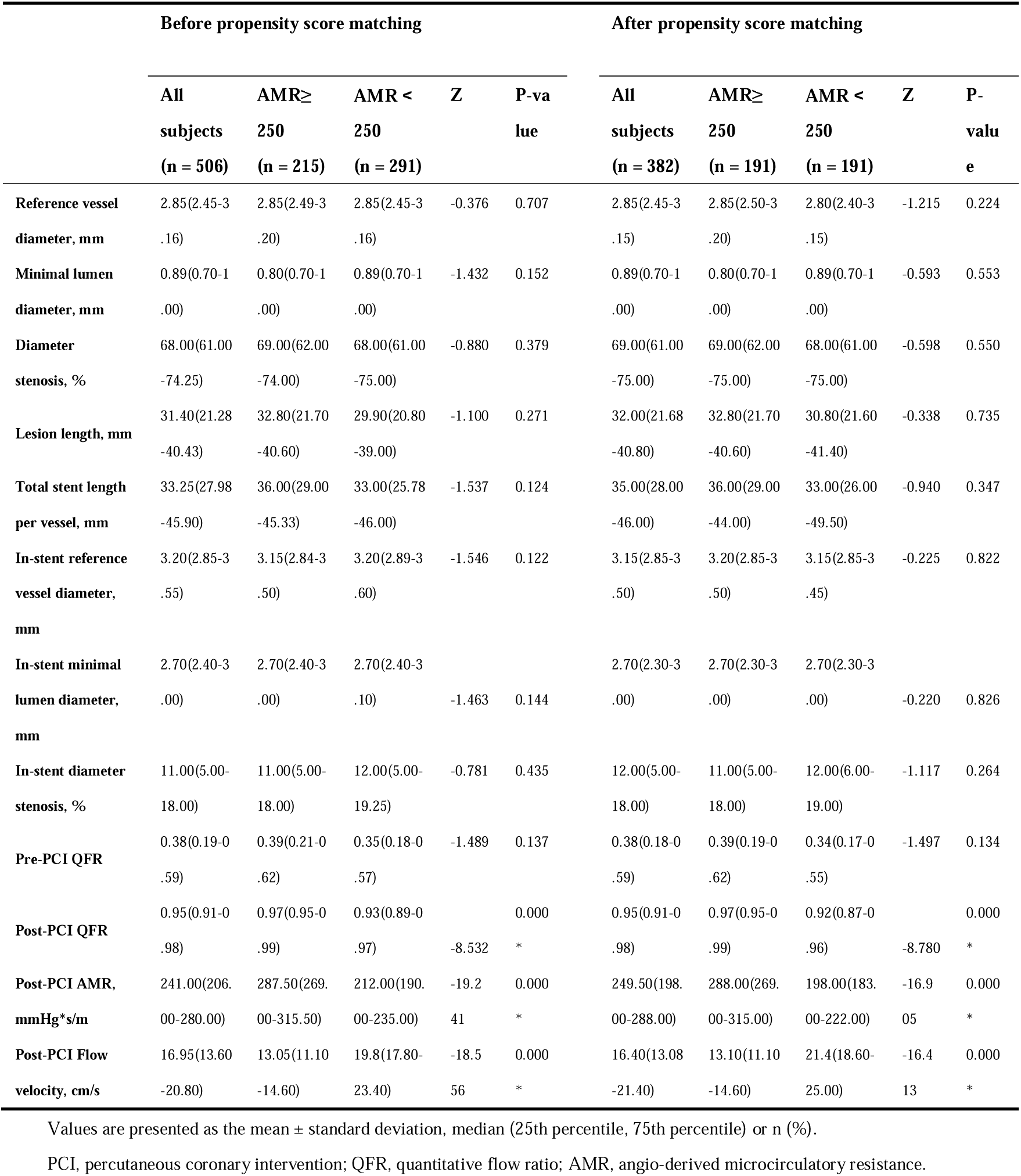
Procedural characteristics

### 3.4 One-Year Follow-Up Characteristics

The clinical characteristics of the two groups at 1 year of follow-up were shown in Table 4. At 1-year follow-up, the primary endpoint occurred in a total of 48 cases (9.5%), of which 30 and 18 cases were from the CMD and Non-CMD groups (HR 2.326 [95% CI 1.297 to 4.172]; 14.0% vs. 6.2%, P=0.005); a total of 22 cases (4.3%) were readmitted for heart failure in the subgroup analysis, of which 16 and 6 cases were from the CMD and Non-CMD groups (HR 3.741 [95% CI 1.464 to 9.562]; 7.4% vs. 2.1%, P=0.006).

**Table 4:**
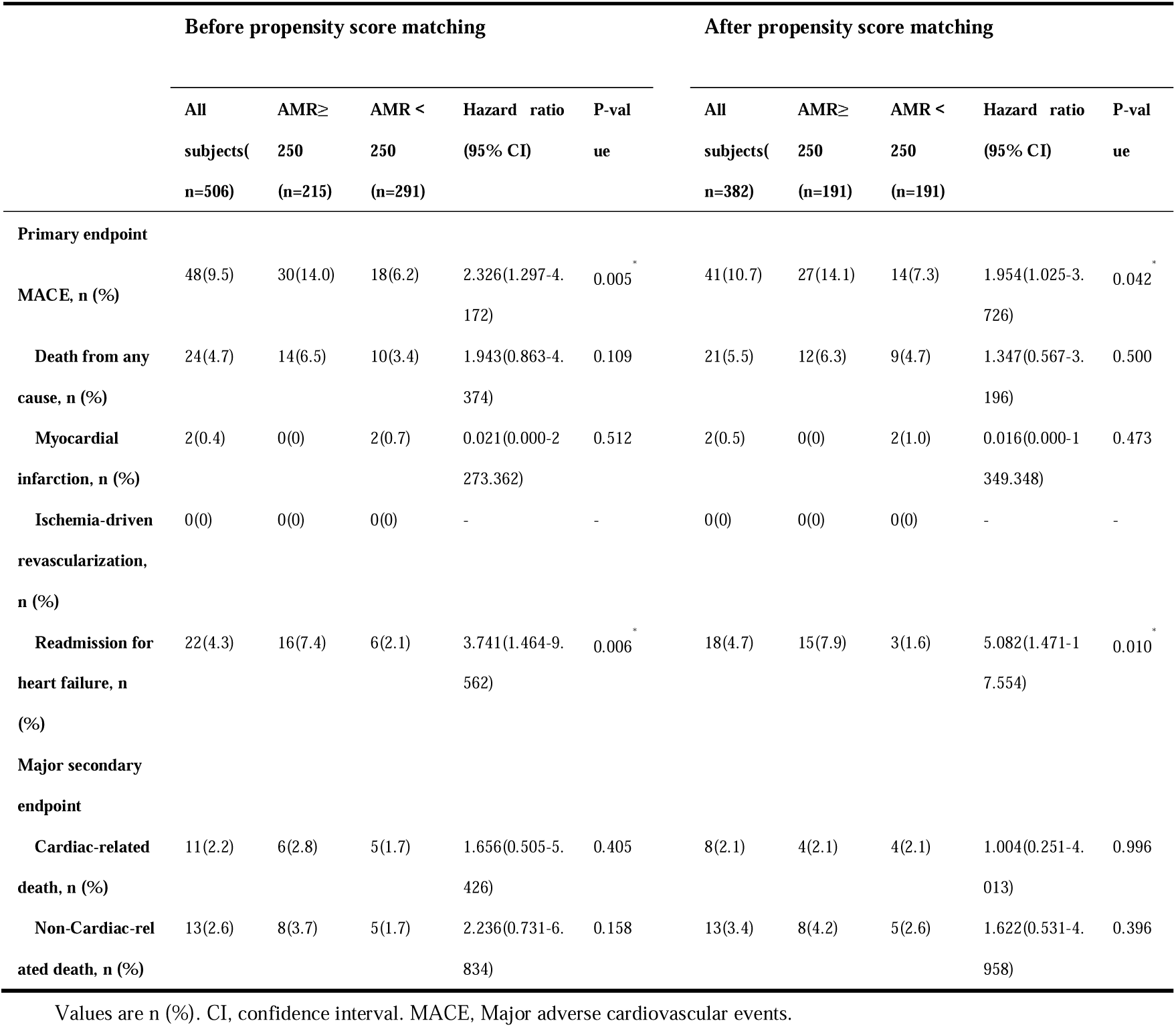
1-year clinical outcomes

Analyses were performed in the intention-to-treat population. Kaplan-Meier curves show the cumulative incidence of MACE, and HF readmission (Figure 3 and Figure 4); while a total of 24 cases (4.8%) had all-cause death, 2 cases (0.4%) had a myocardial infarction, and 0 cases (0%) had target vessel revascularization, and there was no statistical difference between the two groups (P> 0.05). A total of 11 (2.2%) cases of cardiac-related death and 13 (2.6%) cases of non-cardiac-related death occurred in the secondary endpoint, and there was no statistical difference between the two groups for both cardiac-related and non-cardiac-related deaths (P > 0.05).

**Figure 3:**
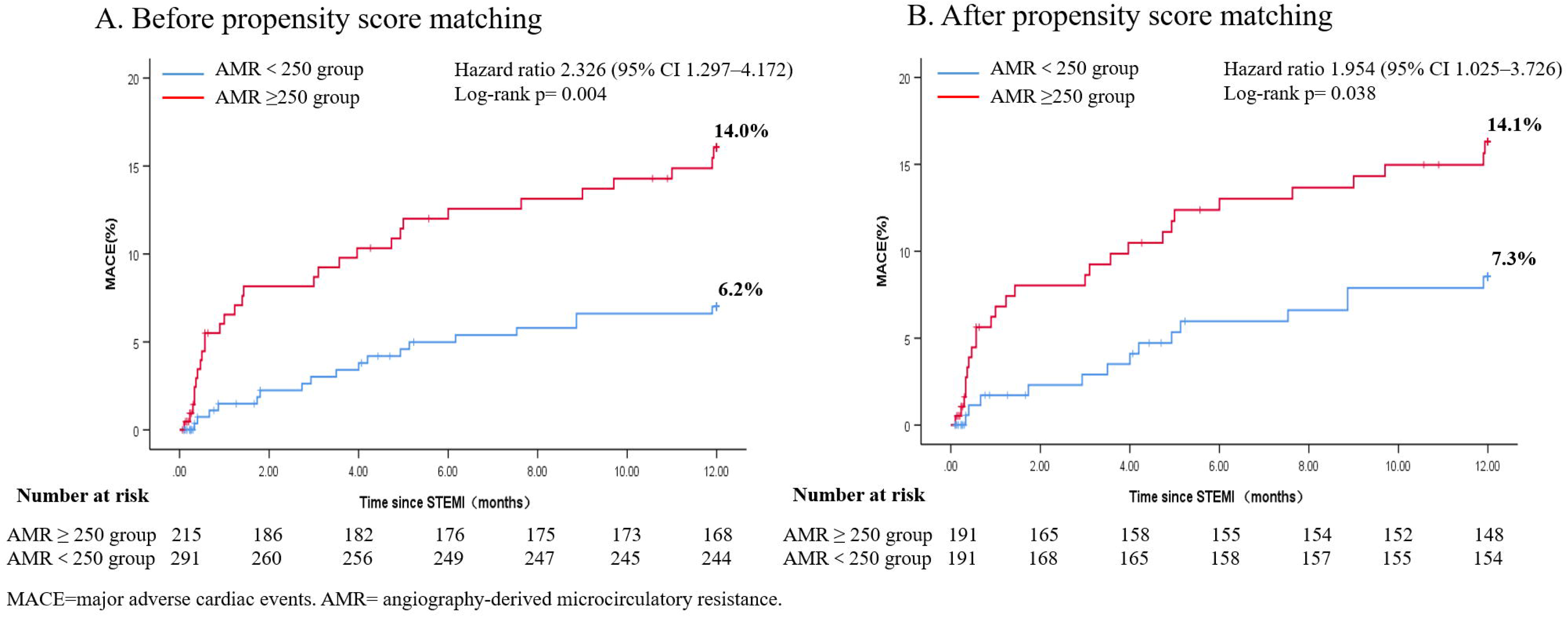
Kaplan-Meier curves for the primary endpoints in the intention-to-treat population

**Figure 4:**
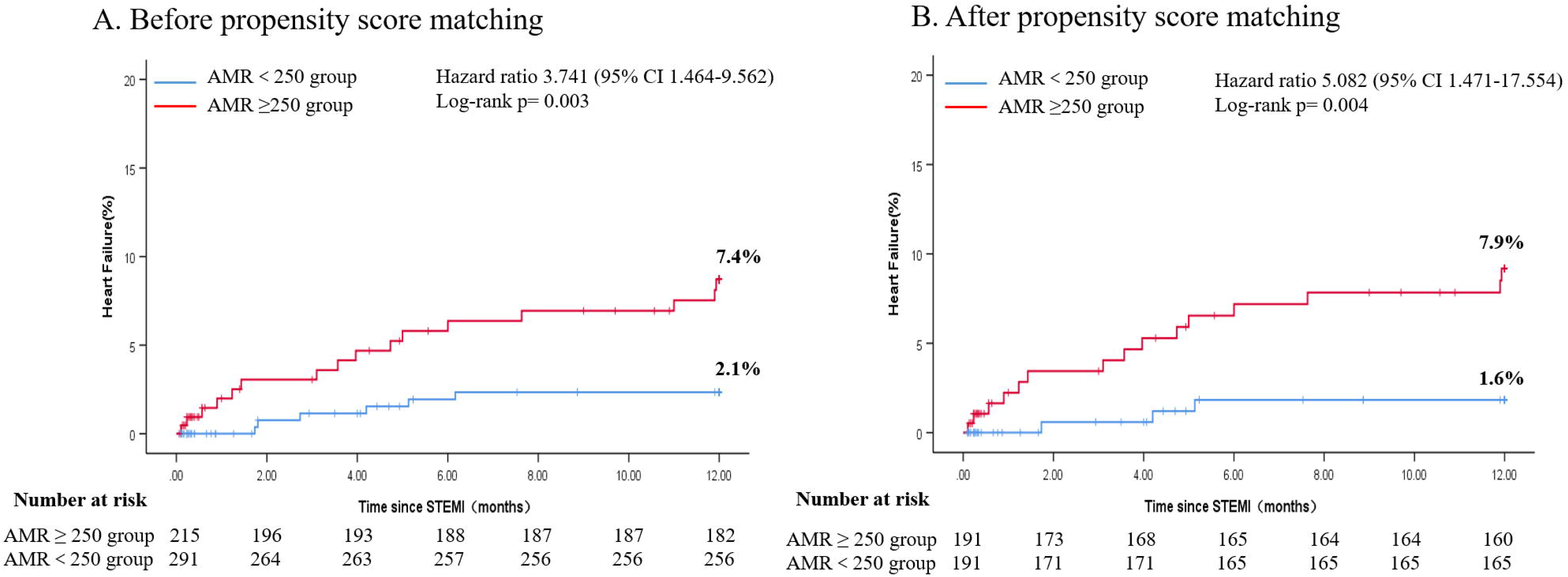
Kaplan-Meier curves for heart failure in the intention-to-treat population

After PSM, at 1-year follow-up, the primary endpoint occurred in a total of 41 cases (10.7%), of which 27 and 14 were from the CMD and Non-CMD groups (HR 1.954 [95% CI 1.025 to 3.726]; 14.1% vs. 7.3%, P=0.042); a total of 18 cases (4.7%) were readmitted for heart failure in the subgroup analysis. Of these, 15 and 3 were from the CMD and Non-CMD groups (HR 5.082 [95% CI 1.471 to 17.554]; 7.9% vs. 1.6%, P=0.010); while a total of 21 (5.5%) had all-cause death, 2 (0.4%) had myocardial infarction, and 0 (0%) had target vessel revascularization, and there was no statistical difference between the two groups (P > 0.05). A total of 8 (2.1%) cases of cardiac-related death and 13 (3.4%) cases of non-cardiac-related death occurred in the secondary endpoint, and there was no statistical difference between the two groups for both cardiac-related and non-cardiac-related deaths (P > 0.05). Contributing to the main composite outcome was the higher incidence of HF readmission by the CMD group compared to the non-CMD group.

Univariate and multifactor predictors of the primary endpoint are listed in Table 5. The 14 independent variables of significance: post-PCI AMR ≥250 mmHg*s/m, age, sex, BMI, diabetes, hypertension, hyperlipidemia, stroke, HBA1c, BNP, Albumin, Peak value of cTnI, random blood glucose, and LVEF were included in the multifactorial COX regression analysis using a univariate COX regression model. Post-PCI AMR ≥250 mmHg*s/m was obtained to be significantly associated with a higher risk of the primary endpoint and was its independent predictor (pre-matched adjusted HR 2.037, 95% CI: 1.068 to 3.888, P = 0.031; post-matched adjusted HR 2.265, 95% CI: 1.136 to 4.515, P = 0.020), as well as age, BNP and Peak value of cTnI (Table 5).

**Table 5:** Univariate and multivariate analysis results for predictors of 1-year MACE

|  | Before propensity score matching |  |  |  |  |  | After propensity score matching |  |  |  |  |  |
| --- | --- | --- | --- | --- | --- | --- | --- | --- | --- | --- | --- | --- |
|  | Univariable analysis |  |  | Multivariable analysis |  |  | Univariable analysis |  |  | Multivariable analysis |  |  |
|  | HR | (95% CI) | p-value | HR | (95% CI) | p-value | HR | (95% CI) | p-value | HR | (95% CI) | p-value |
| **AMR≥250** | 2.326 | (1.297-4.172) | 0.005<sup>*</sup> | 2.037 | (1.068-3.888) | 0.031<sup>*</sup> | 1.954 | (1.025-3.726) | 0.042<sup>*</sup> | 2.265 | (1.136-4.515) | 0.020<sup>*</sup> |
| **Age** | 1.072 | (1.046-1.099) | 0.000<sup>*</sup> | 1.045 | (1.015-1.076) | 0.003<sup>*</sup> | 1.078 | (1.046-1.111) | 0.000<sup>*</sup> | 1.062 | (1.025-1.101) | 0.001<sup>*</sup> |
| **Male** | 1.596 | (0.830-3.068) | 0.161 |  |  |  | 1.030 | (0.476-2.230) | 0.940 |  |  |  |
| **Body mass index** | 0.884 | (0.801-0.975) | 0.014<sup>*</sup> | 1.002 | (0.901-1.114) | 0.972 | 0.898 | (0.807-0.999) | 0.048<sup>*</sup> | 1.010 | (0.901-1.131) | 0.868 |
| **Diabetes mellitus** | 0.308 | (0.122-0.778) | 0.013<sup>*</sup> | 1.119 | (0.298-4.205) | 0.868 | 0.453 | (0.178-1.154) | 0.097 | 1.786 | (0.453-7.042) | 0.407 |
| **Hypertension** | 1.431 | (0.802-2.551) | 0.225 |  |  |  | 1.227 | (0.662-2.274) | 0.515 |  |  |  |
| **Hyperlipidemia** | 0.420 | (0.188-0.936) | 0.034 | 0.692 | (0.302-1.586) | 0.384 | 0.600 | (0.266-1.353) | 0.218 |  |  |  |
| **Previous stroke** | 3.248 | (1.656-6.369) | 0.001<sup>*</sup> | 1.703 | (0.774-3.750) | 0.186 | 3.233 | (1.584-6.597) | 0.001<sup>*</sup> | 1.735 | (0.763-3.948) | 0.189 |
| **HBA1C** | 0.679 | (0.498-0.927) | 0.015<sup>*</sup> | 0.936 | (0.590-1.484) | 0.778 | 0.737 | (0.539-1.008) | 0.056 | 0.928 | (0.586-1.468) | 0.748 |
| **BNP** | 1.000 | (1.000-1.001) | 0.000<sup>*</sup> | 1.000 | (1.000-1.000) | 0.014<sup>*</sup> | 1.000 | (1.000-1.001) | 0.000<sup>*</sup> | 1.000 | (1.000-1.000) | 0.207 |
| **Albumin** | 0.871 | (0.813-0.933) | 0.000<sup>*</sup> | 0.958 | (0.879-1.043) | 0.322 | 0.872 | (0.807-0.942) | 0.000<sup>*</sup> | 0.954 | (0.871-1.044) | 0.305 |
| **Peak troponin I** | 1.017 | (0.999-1.034) | 0.065 | 1.023 | (1.004-1.042) | 0.016<sup>*</sup> | 1.016 | (0.997-1.037) | 0.100 | 1.024 | (1.003-1.046) | 0.024<sup>*</sup> |
| **Glucose** | 0.828 | (0.716-0.958) | 0.011<sup>*</sup> | 0.856 | (0.704-1.040) | 0.118 | 0.851 | (0.735-0.986) | 0.031<sup>*</sup> | 0.818 | (0.659-1.016) | 0.069 |
| **LVEF** | 0.947 | (0.919-0.975) | 0.000<sup>*</sup> | 0.974 | (0.942-1.006) | 0.113 | 0.946 | (0.916-0.976) | 0.000<sup>*</sup> | 0.974 | (0.940-1.009) | 0.137 |

Abbreviations: CI, confidence interval; HR, hazard ratio; other abbreviations as in Tables 1.

## 4. Discussion

In this study, we evaluated CMD in STEMI patients by AMR index, thus exploring the predictive value of AMR on the occurrence of adverse events in STEMI patients. The main findings of this study are as follows:1. Regarding clinical outcomes, we found a higher incidence of MACE in the CMD group, which was mainly due to a higher incidence of heart failure readmission.2. Multivariate COX regression analysis showed that AMR≥250mmHg*s/m, age, peak troponin I and BNP were independent risk factors for 1-year MACE occurrence.

Current diagnostic methods for evaluating CMD include non-invasive tests such as positron emission tomography (PET), cardiac echocardiography, cardiac CT scan, and cardiac magnetic resonance (CMR), as well as invasive tests such as coronary angiography, Doppler flow map, coronary flow reserve (CFR), and IMR. PET is now considered the gold standard reference for non-invasive evaluation of CMD, but its high cost, inability to measure repeatedly, and radiological damage have limited its use in clinical practice [15]; The main limitation of CMR is the adverse reactions caused by gadolinium contrast agent in patients with renal insufficiency. Previously, invasive IMR has been studied more frequently, and research evidence suggests that IMR can accurately assess patients’ CMD [16], and it is now mostly considered that coronary microcirculatory dysfunction is present at IMR ≥ 25 [17]. In previous work, Fearon et al. [18] reported that high IMR > 40 measured at initial PCI in STEMI patients predicted long-term clinical outcomes, such as a death in HF and rehospitalization. IMR of the Culprit vessel measured immediately after successful initial PCI in STEMI patients can assess the prognosis of the patient. However, additional requirements for pressure-temperature transducer wires and insufflators limit their use in routine procedures.

With the increasing variety of functional evaluation, we are no longer limited to the use of assessment tools such as IMR calculated by pressure and temperature lines; Tebaldi M et al. [19] validated for the first time the formula for calculating microvascular resistance based on cQFR data without the use of pressure guidewires and drug-induced congestion. In contrast, a study by Sheng X et al. [20] found that QFR calculation may be a useful tool for predicting CMD after STEMI; as studies have progressed, more and more studies have shown that it is feasible to assess coronary microcirculatory resistance in the absence of intracoronary pressure guidewires and congestive drugs [21]. Studies published during the same period have shown that the non-hyperaemic angiography-derived index of microcirculatory resistance (NH-IMRangio) is prognostically equivalent to invasively measured IMR and may be a viable alternative to IMR in patients with STEMI [22]. In recent studies, the AMR index obtained from a single angiographic view only is a feasible computational alternative to pressure-guided IMR with good diagnostic accuracy in the assessment of CMD [11]. In patients with AMI, current noninvasive examinations and invasive IMR to assess microcirculatory function are less than perfect, which also poses a challenge for early and accurate functional assessment in AMI patients. Undeniably, with the development of functional coronary angiography, we can find AMR to be a good solution to this problem. In STEMI patients, AMR offers a safe and effective, reproducible, and rapidly calculated alternative to guidewire-based IMR measurements.

The prognostic impact of the occurrence of CMD is important for STEMI patients, and previous studies have shown that STEMI patients with Angio-IMR > 40 U have a significantly higher risk of cardiac death or heart failure admission than controls [23], while in a study by Scarsini et al. [16] it was observed that STEMI patients admitted by invasive IMR > 40 U or CMD assessed by CMR showed adverse outcomes mainly caused by the development of heart failure; it is undeniable that PCI significantly reduces in-hospital mortality in STEMI patients, but the incidence of HF after STEMI is not uncommon in clinical practice, which may be that post-ischemic CMD plays an important role and is also consistent with the higher incidence of heart failure readmission in patients in the CMD group in this study results are consistent with this.

CMD is an important pathological change in STEMI patients after reperfusion therapy and is often associated with poor prognosis [24]. Rapid and accurate assessment of CMD in the acute phase of STEMI appears to be important. In a recent study, Yongzhen Fan et al. [11] found that AMR, as a QFR-derived calculated index, had a good correlation (r = 0.83, p < 0.001) and diagnostic performance (AUC 0.94; 95% CI: 0.91 to 0.97) with an optimal threshold value of 250 mmHg *s/m for AMR when IMR ≥ 25. In this study, a subgroup analysis was performed based on this cutoff value, and although there was statistical significance in age, sex ratio, and past medical history between the two groups for the baseline data, the difference between the two baseline data was improved by PSM and made the two groups more comparable, and 14 independent variables that were clinically relevant or significant were included in the multifactor COX regression analysis by univariate COX regression analysis, and the results Post-PCI AMR ≥250 mmHg*s/m after matching was found to be significantly associated with a higher risk of the primary endpoint and was an independent predictor of it.

STEMI is a common acute and critical clinical condition caused by acute myocardial ischemia, prevalent in the elderly population, with rapid progression, and is one of the serious threats to the physical health of the elderly. In our study, baseline data showed that patients with CMD were significantly older compared to patients with the non-CMD [67.00 (56.00-74.00) vs. 60.00 (51.75-72.00) years, P=0.000]; after incorporating multifactorial COX regression analysis, it was concluded that age in STEMI patients was associated with a higher risk of the primary endpoint association (adjusted HR 1.045, 95% CI: 1.015-1.076, P = 0.003).

BNP is a counter-regulatory peptide hormone synthesized mainly in the ventricular myocardium. As a highly sensitive and specific biomarker of the degree of myocardial infarction, in non-ST-segment elevation ACS patients, patients with high BNP at presentation are at higher risk of death and congestive heart failure[25]and is an independent predictor of very long-term all-cause mortality [26]. Elevated BNP concentrations at initial presentation in STEMI patients are associated with a higher risk of death in the short term [27] [28]. And in our study, we also came to the consistent conclusion that BNP was higher in CMD patients compared to the non-CMD patients [189.19 (78.09-485.36) vs. 142.55 (50-401.82) pg/ml, P=0.011], and after including a multifactorial COX regression analysis, it was concluded that high BNP at admission in STEMI patients was associated with a major higher risk at the endpoint (adjusted HR 1.000, 95% CI: 1.000-1.000, P = 0.019).

In patients with NSTE-ACS treated with PCI, higher peak pre-procedure cTnI levels were independently associated with 30-day mortality and composite MACE [29]. By 3-month follow-up, troponin I was associated with clinical outcomes and cardiac function in STEMI patients treated with initial PCI [30]. Whereas in our findings, neither troponin I nor peak troponin I were statistically different in CMD patients admitted to the hospital compared to the non-CMD patients (P > 0.05), after inclusion of multifactorial COX regression analysis, it was concluded that peak troponin I in STEMI patients was associated with a higher risk of the primary endpoint (adjusted HR 1.022, 95% CI: 1.004 -1.041, P= 0.019).

Diabetes mellitus is a metabolic disease with macrovascular and microvascular complications. Since endothelial dysfunction plays an important role in the cardiovascular complications of diabetes, an association between previous history of diabetes, hyperglycemia, and post-ischemic CMD seems reasonable. A prospective trial of non-diabetic patients presenting for the first time with STEMI confirmed that hyperglycemia on admission was significantly associated with MVO as defined by CMR [31]. In contrast, in our study, history of diabetes, glucose level at admission, and HbA1c were not statistically significant between the CMD and Non-CMD groups, nor was the inclusion of HbA1c in the multifactorial COX regression analysis between groups (adjusted HR 0.932, 95% CI: 0.587 to 1.480, P= 0.766); this may be related to our lower prevalence of diabetes mellitus in the included population and the smaller sample size.

Recent advances in functional coronary angiography have made it possible to estimate physiologic indicators based on coronary angiography alone, without the use of pressure-temperature sensor wires or congestive drugs. In the present study, we evaluated the prediction of adverse events by AMR in patients with STEMI. In STEMI patients, early assessment of AMR is convenient and allows earlier administration of medications for microcirculatory disorders. QFR and AMR may be more readily adopted into the workflow of angiography-based diagnostic and interventional procedures than invasive physiologic assessments, do not require the use of specialized guidewires, and can be easily repeated multiple times during the procedure. It may thus facilitate the routine use of physiological assessment in clinical practice.CMD is prevalent in patients with cardiovascular risk factors and is associated with an increased risk of adverse events and is an important cause of coronary artery disease (CAD) [2]. Some investigators have suggested that oxidative stress and inflammatory responses caused by excessive production and accumulation of cellular reactive oxygen species (ROS) are the key mechanisms driving the development of CMD [32]. In vitro and in vivo studies have shown that increased intracellular ROS concentrations promote the conversion of NO in peroxynitrite radicals, leading to impaired NO-mediated vasodilation and enhanced vasoconstrictor activity of ET-1 (vasoconstrictor agonist) through activation of the RhoA/Rho-kinase pathway [33] [34]. In patients with STEMI, there is no effective treatment to improve MVO [35]; no large-scale randomized clinical trials have been seen to study specific treatment strategies for CMD, the pathophysiological mechanisms of CMD in different cardiovascular diseases have not been fully elucidated, and there is a lack of treatment options for CMD, which require further in-depth studies to achieve the goal of providing individualized treatment for patients [2]. Therefore, active control of risk factors including smoking cessation, rational control of blood pressure and diabetes, lipid management, and treatment of the primary disease are effective means to prevent the progression of microangiopathy and improve angina symptoms. In addition, early identification of high-risk patients is crucial as early development of individualized treatment strategies to improve long-term prognosis.

The present study still has some limitations. First, this is a retrospective single-center observational study with a small sample size, and further prospective multicenter cohort studies are needed to validate the findings. Second, not all images are suitable for QFR and AMR analysis, which may lead to selection bias. Third, individual vessel AMR immediately after primary PCI may not fully explain the overall prognosis of patients, and the focus of this study was limited to the analysis of microcirculatory dysfunction in the vascular region of the culprit in STEMI patients. Therefore, studies such as comparative prognosis of UA, NSTEMI patients, and non-culprit vascular regions with potential microcirculatory dysfunction may be the next research plan of our group. Fourth, the MACE in this study was a 1-year follow-up, which is a short period, and the follow-up will continue for a longer period. In addition, since most of the follow-ups were conducted by telephone and returned to the hospital, which was affected by economic conditions and epidemics, it is a pity that there is no data on coronary reexamination 1 year after myocardial infarction.

## 5. Conclusion

With the development of techniques and devices, AMR as a guidewire-free and adenosine-free index for the assessment of coronary microcirculatory impairment may become a viable alternative to invasive guidewire-based IMR in patients with STEMI.

## Data Availability

All data produced in the present study are available upon reasonable request to the authors

## References

1. Crea F, Camici PG, Bairey Merz CN. Coronary microvascular dysfunction: an update. Eur Heart J. 2014;35:1101–1111.

2. Del Buono MG, Montone RA, Camilli M, Carbone S, Narula J, Lavie CJ, Niccoli G, Crea F. Coronary Microvascular Dysfunction Across the Spectrum of Cardiovascular Diseases: JACC State-of-the-Art Review. J Am Coll Cardiol. 2021 Sep 28;78(13):1352–1371.

3. Niccoli G, Montone RA, Ibanez B, et al. Optimized Treatment of ST-Elevation Myocardial Infarction. Circ Res. 2019;125(2):245–258.

4. Fearon WF, Balsam LB, Farouque HM, et al. Novel index for invasively assessing the coronary microcirculation. Circulation. 2003;107:3129–3132.

5. Tu S, Westra J, Yang J, et al. Diagnostic accuracy of fast computational approaches to derive fractional flow reserve from diagnostic coronary angiography: the international multicenter FAVOR pilot study. JACC Cardiovasc Interv 2016; 9: 2024–2035.

6. Westra J, Andersen BK, Campo G, et al. Diagnostic performance of inDprocedure angiographyDderived quantitative flow reserve compared to pressureDderived fractional flow reserve: the FAVOR II EuropeDJapan study. J Am Heart Assoc 2018; 7: e009603.

7. Xu BO, Tu S, Qiao S, et al. Diagnostic accuracy of angiographyDbased quantitative flow ratio measurements for online assessment of coronary stenosis. J Am Coll Cardiol 2017; 70: 3077–3087.

8. Westra J, Tu S, Winther S, et al. Evaluation of coronary artery stenosis by quantitative flow ratio during invasive coronary angiography: the WIFI II study (WireDFree Functional Imaging II). Circ Cardiovasc Imaging 2018; 11: e007107.

9. Spitaleri G, Tebaldi M, Biscaglia S, et al. Quantitative flow ratio identifies nonculprit coronary lesions requiring revascularization in patients with STDsegmentDelevation myocardial infarction and multivessel disease. Circ Cardiovasc Interv 2018; 11: e006023.

10. Xu BO, Tu S, Song L, et al. Angiographic quantitative flow ratioDguided coronary intervention (FAVOR III China): a multicentre, randomised, shamDcontrolled trial. Lancet 2021; 398: 2149–2159.

11. Fan, Y, Fezzi S, Sun P, et al. In Vivo Validation of a Novel Computational Approach to Assess Microcirculatory Resistance Based on a Single Angiographic View. Journal of Personalized Medicine 12.11 (2022): 1798.

12. Ibanez, B.; James, S.; Agewall, S.; Antunes, M.J.; Bucciarelli-Ducci, C.; Bueno, H.; Caforio, A.L.P.; Crea, F.; Goudevenos, J.A.; Halvorsen, S.;, et al. 2017 ESC Guidelines for the management of acute myocardial infarction in patients presenting with STsegment elevation: The Task Force for the management of acute myocardial infarction in patients presenting with ST-segment elevation of the European Society of Cardiology (ESC). Eur. Heart J. 2018, 39, 119–177.

13. Tu S, Echavarria-Pinto M, von Birgelen C, et al. Fractional flow reserve and coronary bifurcation anatomy: a novel quantitative model to assess and report the stenosis severity of bifurcation lesions. JACC Cardiovasc Interv. 2015;8:564–574.

14. Thygesen K, Alpert JS, Jaffe AS, Simoons ML, Chaitman BR, White HD, et al. Third universal definition of myocardial infarction. Circulation. (2012) 126:2020–35.

15. Ong P, Safdar B, Seitz A, Hubert A, Beltrame JF, Prescott E. Diagnosis of coronary microvascular dysfunction in the clinic. Cardiovasc Res. 2020;116(4):841–855.

16. Scarsini R, Shanmuganathan M, De Maria GL, Borlotti A, Kotronias RA, Burrage MK, Terentes-Printzios D, Langrish J, Lucking A, Fahrni G, Cuculi F, Ribichini F, Choudhury RP, Kharbanda R, Ferreira VM, Channon KM, Banning AP; OxAMI Study Investigators. Coronary Microvascular Dysfunction Assessed by Pressure Wire and CMR After STEMI Predicts Long-Term Outcomes. JACC Cardiovasc Imaging. 2021 Oct;14(10):1948–1959.

17. Fearon WF, Kobayashi Y. Invasive Assessment of the Coronary Microvasculature: The Index of Microcirculatory Resistance[J]. Circ Cardiovasc Interv, 2017, 10(12): e005361.

18. Fearon WF, Low AF, Yong AS, et al. Prognostic value of the Index of Microcirculatory Resistance measured after primary percutaneous coronary intervention. Circulation. 2013;127(24):2436–2441.

19. Tebaldi M, Biscaglia S, Di Girolamo D, Erriquez A, Penzo C, Tumscitz C, Campo G. Angio-Based Index of Microcirculatory Resistance for the Assessment of the Coronary Resistance: A Proof of Concept Study. J Interv Cardiol. 2020 Oct 25;2020:8887369.

20. Sheng X, Qiao Z, Ge H, Sun J, He J, Li Z, Ding S, Pu J. Novel application of quantitative flow ratio for predicting microvascular dysfunction after ST-segment-elevation myocardial infarction. Catheter Cardiovasc Interv. 2020 Feb;95 Suppl 1:624–632.

21. Mejia-Renteria H, Lee JM, Choi KH, Lee SH, Wang L, Kakuta T, Koo BK, Escaned J. Coronary microcirculation assessment using functional angiography: Development of a wire-free method applicable to conventional coronary angiograms. Catheter Cardiovasc Interv. 2021 Nov 15;98(6):1027–1037.

22. Kotronias RA, Terentes-Printzios D, Shanmuganathan M, Marin F, Scarsini R, Bradley-Watson J, Langrish JP, Lucking AJ, Choudhury R, Kharbanda RK, Garcia-Garcia HM, Channon KM, Banning AP, De Maria GL. Long-Term Clinical Outcomes in Patients With an Acute ST-Segment-Elevation Myocardial Infarction Stratified by Angiography-Derived Index of Microcirculatory Resistance. Front Cardiovasc Med. 2021 Sep 7;8:717114.

23. Choi KH, Dai N, Li Y, Kim J, Shin D, Lee SH, Joh HS, Kim HK, Jeon KH, Ha SJ, Kim SM, Jang MJ, Park TK, Yang JH, Song YB, Hahn JY, Doh JH, Shin ES, Choi SH, Gwon HC, Lee JM. Functional Coronary Angiography-Derived Index of Microcirculatory Resistance in Patients With ST-Segment Elevation Myocardial Infarction. JACC Cardiovasc Interv. 2021 Aug 9;14(15):1670–1684.

24. Niccoli G, Burzotta F, Galiuto L, Crea F. Myocardial no-reflow in humans. J Am Coll Cardiol. 2009;54:281–292.

25. Morrow DA, de Lemos JA, Sabatine MS, Murphy SA, Demopoulos LA, DiBattiste PM, McCabe CH, Gibson CM, Cannon CP, Braunwald E. Evaluation of B-type natriuretic peptide for risk assessment in unstable angina/non-ST-elevation myocardial infarction: B-type natriuretic peptide and prognosis in TACTICS-TIMI 18. J Am Coll Cardiol. 2003 Apr 16;41(8):1264–72.

26. Bassan F, Bassan R, Esporcatte R, Santos B, Tura B. Very Long-Term Prognostic Role of Admission BNP in Non-ST Segment Elevation Acute Coronary Syndrome. Arq Bras Cardiol. 2016 Mar;106(3):218–25.

27. Mega Jessica L, Morrow David A, De Lemos James A et al. B-type natriuretic peptide at presentation and prognosis in patients with ST-segment elevation myocardial infarction: an ENTIRE-TIMI-23 substudy.[J]. J Am Coll Cardiol, 2004, 44: 335–9.

28. Grabowski M, Filipiak KJ, Karpinski G, Wretowski D, Rdzanek A, Rudzki D, Główczyńska R, Rudowski R, Opolski G. Prognostic value of B-type natriuretic peptide levels on admission in patients with acute ST-elevation myocardial infarction. Acta Cardiol. 2005 Oct;60(5):537–42.

29. Napan S, Kashinath RC, Kadri S, Orig MN, Khadra S. Prognostic significance of preprocedural troponin-I in patients with non-ST elevation acute coronary syndromes undergoing percutaneous coronary intervention. Coron Artery Dis. 2010 Aug;21(5):261–5.

30. Hall TS, Hallén J, Krucoff MW, Roe MT, Brennan DM, Agewall S, Atar D, Lincoff AM. Cardiac troponin I for prediction of clinical outcomes and cardiac function through 3-month follow-up after primary percutaneous coronary intervention for ST-segment elevation myocardial infarction. Am Heart J. 2015 Feb;169(2):257–265.

31. Jensen CJ, Eberle HC, Nassenstein K, Schlosser T, Farazandeh M, Naber CK, Sabin GV, Bruder O. Impact of hyperglycemia at admission in patients with acute ST-segment elevation myocardial infarction as assessed by contrast-enhanced MRI. Clin Res Cardiol 2011;100:649–659.

32. Masi S, Rizzoni D, Taddei S, et al. Assessment and pathophysiology of microvascular disease: recent progress and clinical implications. Eur Heart J. 2020;ehaa857.

33. Magenta A, Greco S, Capogrossi MC, Gaetano C, Martelli F. Nitric oxide, oxidative stress, and p66Shc interplay in diabetic endothelial dysfunction. Biomed Res Int. 2014;2014:193095.

34. Tsai SH, Lu G, Xu X, Ren Y, Hein TW, Kuo L. Enhanced endothelin-1/Rho-kinase signaling and coronary microvascular dysfunction in hypertensive myocardial hypertrophy. Cardiovasc Res. 2017;113(11):1329–1337.

35. Tomanek RJ. Structure–Function of the Coronary Hierarchy: Coronary Vasculature: New York: Springer; 2013:59–81.

